# Impact of universal masking in health care and community on SARS-CoV-2 spread

**DOI:** 10.1101/2020.09.02.20187021

**Authors:** MW Pletz, A Steiner, M Kesselmeier, B Löffler, S Trommer, S Weis, J Maschmann, A Stallmach

## Abstract

Universal masking the health care setting and in the community to contain the spread of SARS-CoV-2 has been recently recommended by the WHO, but supporting data are rare. The City of Jena was the first community in Germany to issue an order on mandatory public masking. Here, we report the development of the number of novel infections in our hospital and in the city of Jena after implementation of universal masking in our hospital and the city.

## Introduction

There are only a few studies on universal masking during the flu season in hospitals (Esposito et al. 2020), and, to the best of our knowledge, there are no studies investigating the efficacy of universal masking in healthy individuals to prevent SARS-CoV-2 transmissions. At least, for seasonal human corona virus infection, it was shown that exhaled droplets with a diameter of ≤5 µm containing viral particles can completely be withheld by surgical masks when worn by an infected person (Leung et al. 2020). Arguments against universal masking include a global supply shortage of medical masks, the insufficient filtration of alternative cloth masks compared to medical masks (MacIntyre et al. 2015), a false sense of protection by wearing a mask that may result in failing to maintain physical distance from others, and the risk imposed by incorrect usage of masks by non-trained carriers, *e.g*. transmitting pathogens from contaminated hands to the mask. Here, we report effects of universal masking that were observed after implementation of (*i*) mandatory universal masking in the Jena University Hospital (JUH) and (*ii*) of mandatory community masking in the city of Jena.

## Impact of hospital-wide universal masking

The JUH is a tertiary academic hospital with about 1,400 beds and 5,600 employees providing care for about 53,000 inpatients per year. The first detected COVID-19 case in Jena was a physician returning from Austria. Upon his return, he immediately reported to the hospital occupational health department, was tested and isolated at home on 11^th^ March 2020. On 16^th^ March, a nurse that had returned from skiing in Northern Italy became the source of the first nosocomial outbreak involving four patients and two health care workers (HCW). Consequently, extensive screening was conducted on 1,311 HCW – representing one third of the total work force in direct patient care-between 11^th^ March and 12^th^ May resulting in 31 positive cases until 19^th^ March, approximately one week after the first confirmed case (***Figure 1***). As a consequence, mandatory masking for all personnel involved in patient care was implemented on 20^th^ March. Continued routine staff screening revealed only four HCWs tested positive between 20^th^ March and 12^th^ May. This corresponds to a substantial drop in the rate of new infections among HCWs from 10.1% (31 of 306 screened HCW in this period) before to 0.4% (4 of 1,005 HCW screened in this period) after implementation of universal masking. In addition, a strict containment system was introduced on 5^th^ April, which included placing all patients in single rooms upon admission and SARS-CoV-2 screening regardless of their admitting chief complaint. Thereafter, HCW screening did not reveal any further positive case.

**Figure 1:**
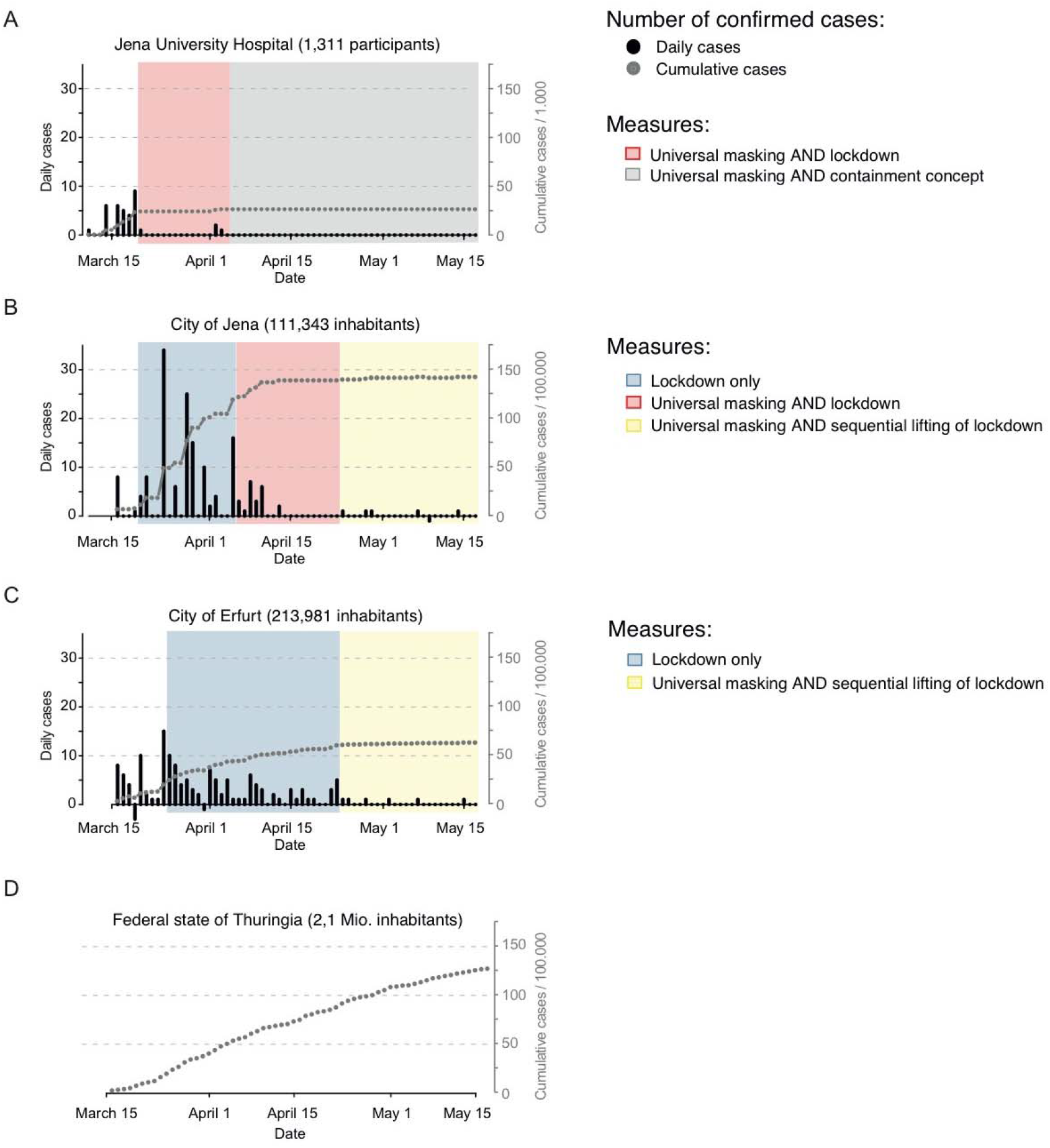
Daily reported newly confirmed cases of SARS-CoV-2 infection and **A)** corresponding cumulative cases per 1,000 health care workers (HCW) of Jena University Hospital (JUH, 1,500 beds as well as per 100,000 inhabitants of the Thuringian cities **B)** Jena and **C)** Erfurt. All reported cases comprise only the first SARS-CoV-2 test results. Results of multiple testing are excluded because all infected subjects were reported by full name and address to the respective Public Health Departments. **D)** For comparison, the cumulative number per 100,000 inhabitants in the State of Thuringia is provided. Implemented measures are color-coded. Of note, “negative” bars for daily reported novel infections in Jena and Erfurt are explained by re-classification of individual COVID-19 cases as non-COVID-19 after quality control of the respective data and laboratory reports by the State Health Department of Thuringia. The Population was taken from the Thuringian State Office of Statistics (31st December 2019).

## Impact of community-wide universal masking

Jena is the second largest city in Thuringia, Germany, with approximately 111,400 inhabitants. Despite thorough contact tracing and isolation of confirmed COVID-19 positive cases by the authorities, a rapid increase in numbers was observed after patient zero (***Figure 1***). The governmental containment (closing of stores, schools, kindergartens, churches etc.) together with social distancing measures was strictly implemented on 20^th^ March in Jena and four days later in the entire Federal State of Thuringia. As novel infections continued to occur, Jena authorities were the first in Germany to introduce universal community-wide masking*, i.e*. mandatory covering of the mouth and nose in public buildings and public transport accepting also cloth masks or scarfs. On 24^th^ April, a state-wide recommendation for community-wide masking in combination with first relaxations of the strict lockdown was issued.

We also assessed the effectiveness of mandatory community-wide masking in Jena in comparison with the nearby city of Erfurt (***Figure 1***). Both cities exhibit a similar university as well as socioeconomic structure. Erfurt introduced masking four weeks after Jena, simultaneously with the first relaxation of the lockdown measures.

As shown in ***Figure 1***, there were no new COIVD-19 cases in Jena five days after implementation, which is in line with the average incubation time of SARS-CoV-2. While in Erfurt, COVID-19 continued to spread and only ceased after the same community-wide masking was imposed.

We are aware that mere association is not causation and that our conclusions are limited by the observational nature of the data. However, it is uncertain whether an ethically sound trial on the controversial issue of mandatory community-wide masking will ever be performed. Therefore, our observations support the notion to implement universal masking in both health care as well as community settings as considering the ensuing reduced infection-rates. Given the risk-benefit ratio, we consider universal masking combined with social distancing as a suitable measure to contain the spread of SARS-CoV-2.

## Data Availability

The data that support the findings of this study are available from the corresponding author, [author initials], upon reasonable request.

